# Associations between sedentary behaviour and motor competence in 3–4-year-olds: A Systematic review

**DOI:** 10.1101/2023.11.01.23297895

**Authors:** Nana A Kwofie, Xanne Janssen, John J Reilly

## Abstract

**BACKGROUND:** Several studies have reported low adherence to World Health Organization sedentary behaviour guidelines in the early years. The purpose of this review is to test for associations between time spent in different types of sedentary behaviour (screen time, habitual sedentary behaviour) and motor competence (fundamental motor skills, fine and gross motor skills, locomotor skills, object control and balance) in 3–4-year-olds.

**METHODS:** Five databases were searched on the 27^th^ of July 2021 with an updated search conducted on the 30^th^ of September 2023: Web of Science (core collection), PUBMED Central, EMBASE (Ovid), SPORT Discus and ERIC. Studies were included in the review if they reported on an association between time spent in sedentary behaviour at ages 3-4 years and motor competence. The methodological quality for each of the included studies was assessed using Joanna Briggs Institute critical appraisal tools. Vote counting was used to determine the direction of associations.

**RESULTS:** Of 5276 total studies found in the search, 16 studies (12 cross-sectional, 4 longitudinal) from 11 different countries met the inclusion criteria. Seven studies examined the association between screen time and motor competence, six examined associations between time spent in sedentary behaviour with motor competence, and three papers examined the association of both screen time and time spent in sedentary behaviour with motor competence. Vote counting showed the direction of association to be predominantly negative for both screen time and time spent in habitual sedentary behaviour with the different components of motor competence. Quality of evidence ranged from 3-7 out of 9 for cross-sectional studies and 6-9 out of 12 for longitudinal studies.

**CONCLUSION:** There may be negative associations between time spent sedentary and motor competence in 3–4-year-olds. However, future studies with stronger study design are required to confirm these associations. Findings from this review should be considered when designing strategies and interventions to promote adherence to sedentary behaviour guidelines.

## Background

The World Health Organisation (WHO) has identified the early years as a critical time to intervene to improve child health and development, and highlights physical activity, sleep and sedentary behaviour (SB) as key behaviours which should be targeted^1,2^ For SB, WHO recommends no more than 60 minutes of sedentary screen time and not being restrained for more than an hour at a time in a 24-hour period for 3–4-year olds^1^. Some studies have shown levels of screen time among young children to be typically more than 3 hours a day, especially during the Covid-19 pandemic lockdown period^3,4^. High levels of SB may negatively impact a range of health outcomes including motor competence (MC)^5^. Motor competence, which is an individual’s degree of proficient performance in a broad range of motor skills, is a crucial part of a young child’s development and has been considered as a foundation for physical activity in young children^6^. Under the umbrella of motor competence, different terminologies such as fundamental motor or movement skills, motor proficiency, motor coordination, motor ability, fine and gross motor skills have been used in the literature^6^. One misconception is that children will attain normal levels of MC naturally as they get older, but many children do not^7^. Considering the importance of developing MC in children, it is therefore important to examine SB as a factor that can potentially affect MC in early childhood.

Poitras et al.^8^ reviewed the relationship between SB and health indicators including MC in the early years. Studies up until April 2016 were included and results from that review showed null associations with SB and MC - only 7 eligible papers were identified and the quality of evidence of the included studies was also very low. However, since the publication of that review, there has been an increased interest in research in the area of SB with more publications emerging. Hence an update on the review by Poitras et al.^8^ to look at more recent evidence and examine associations will be timely. A more recent systematic review by Dos Santos et al.^9^ found negative associations between time spent in SB and MC. However, this review did not examine the association between different types of SB and MC separately and did not analyse individual MC subgroups such as locomotor skills, object control and balance. Dos Santos et al.^9^ also considered children aged 3-6 years as a single category which is not in line with the age group of the 24hr movement guidelines by the WHO (3-4 years)^2^.

An updated review is therefore required to investigate relationships between different types of SB (screen time, habitual SB) and different components of MC among 3- and 4-year-olds. Therefore, the aim of this study is to examine associations between both screen time and habitual SB, and their association with different components of MC (fundamental motor skills, fine and gross motor skills, locomotor skills, object control and balance) in 3-to 4-year-olds.

## Methods

### Protocol and registration

This systematic review was registered and reported by following the Preferred Reporting Item for Systematic Reviews (PRISMA) guideline. This systematic review is registered with the International Prospective Register of Systematic Reviews PROSPERO, with the registration no CRD42021253381.

### Study inclusion criteria

This systematic review used the PECO framework (population, exposure, comparisons, and outcome) to identify key study concepts for the research question (See table 1).

**Table 1.**
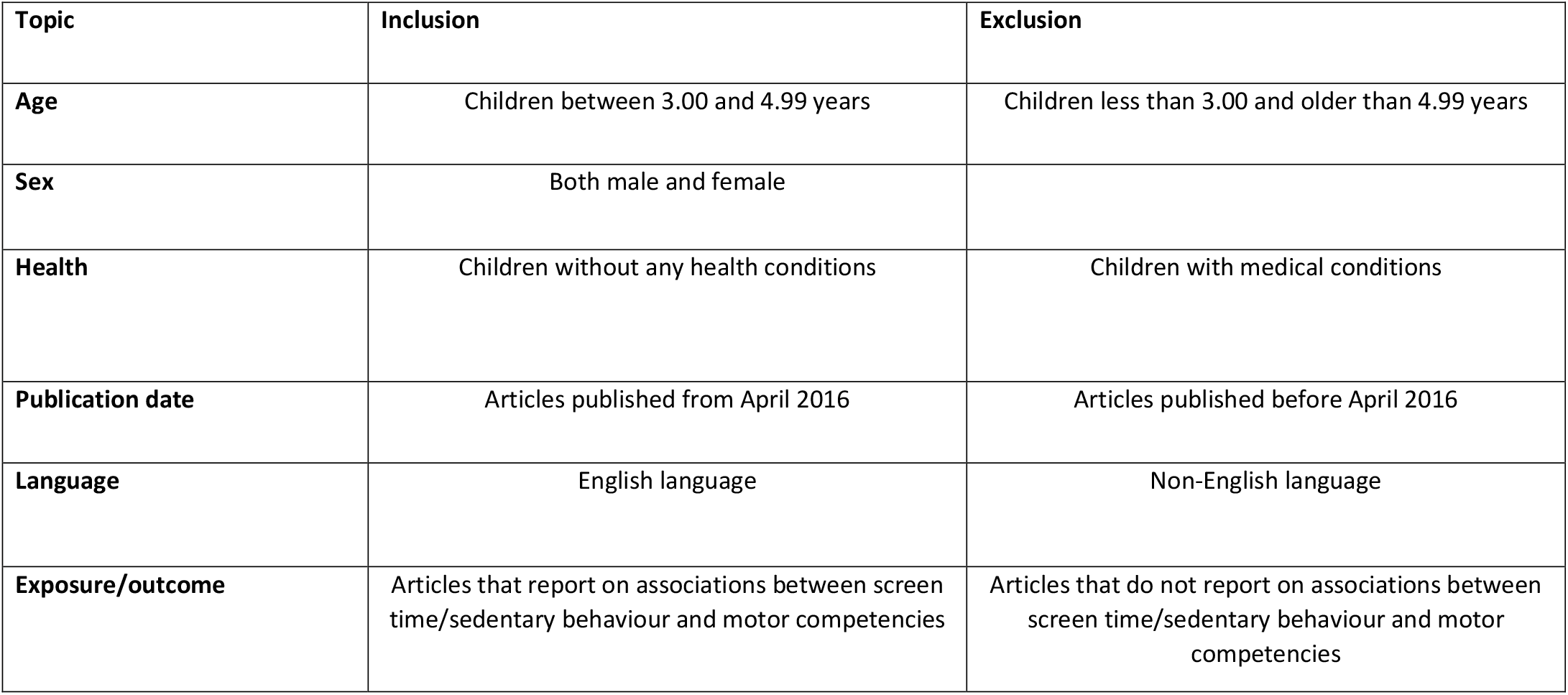
Inclusion and exclusion criteria table.

| **Topic** | **Inclusion** | **Exclusion** |
| --- | --- | --- |
| **Age** | Children between 3.00 and 4.99 years | Children less than 3.00 and older than 4.99 years |
| **Sex** | Both male and female |  |
| **Health** | Children without any health conditions | Children with medical conditions |
| **Publication date** | Articles published from April 2016 | Articles published before April 2016 |
| **Language** | English language | Non-English language |
| **Exposure/outcome** | Articles that report on associations between screen time/sedentary behaviour and motor competencies | Articles that do not report on associations between screen time/sedentary behaviour and motor competencies |

### Population

Articles reporting on children who were considered healthy between the ages of 3.0 and 4.9 years were included in this review. This age group definition is in line with the WHO guidelines on the 24hr movement behaviours for 3-4.9-year olds^2^. Articles that reported on children between the ages of 3 and 4.9 years with diseases or existing health conditions that could potentially affect the outcome were excluded from the review. Also, articles which reported on a sample of children with a mean age below 3.0 or above 4.9 years were excluded unless sub-analysis results were provided for those 3.0-4.9 years of age.

### Exposure

The exposure was a measure of usual (habitual) time spent sedentary and/or on a screen (e.g., television time, time spent on mobile devices, sitting time, video gaming). Different types of screen time were classified as one category for this review. Studies that used either objective (e.g., accelerometer) and/or subjective measures (e.g., parent questionnaire, diaries) were included. For this review, SB was defined as any waking behaviour characterized by an energy expenditure of ≤1.5 METs while in a sitting or reclining posture^10^. Studies that defined SB based on low levels of physical activity, such as ‘physical inactivity’ were not included.

### Comparison

If studies reported on various durations, interruptions, pattern, and types of sedentary behaviour, these were used for comparison.

### Outcome

The primary health outcome for this review is MC which includes locomotor skills (such as jumping, running sliding, galloping), object control (such as striking and kicking) and balance (non-locomotor skills that focus on balance)^6^. Studies that measured any form of motor competence as an outcome with the use of objective measuring such as the Ages and Stages questionnaire were included. Cross-sectional, longitudinal, intervention or experimental study designs that met the above inclusion criteria and were published in English language after April 2016 were included in the review.

### Information sources and search strategy

An electronic literature search was conducted on the following databases on the 27^th^ of July 2021, (with an updated search conducted on 30^th^ September 2023): Web of Science core collection, PUBMED Central, EMBASE (Ovid), SPORT Discuss and ERIC. The search period was set between April 2016 and September 2022. The search period started from when the previous review by Poitras^8^ ended. The full search strategy can be found in Appendix A. Example key words used for the population search included early childhood, preschool children; for the exposure search, sedentary behaviour, screen time, sitting time, and for the outcome search, motor competence, fine motor skill, gross motor skill. Search results for each database were exported to EndNote Reference manager (Version X9), and duplicates were removed.

### Data screening

NK and XJ independently screened the titles and abstract of all articles. If articles were deemed to meet the inclusion criteria full text copies were obtained. Full text screening was performed independently by the same reviewers. In the case of conflicts between two reviewers, inclusion was decided based on discussion between reviewers or if needed, a third reviewer (JJR) was consulted.

### Data extraction

Data extraction was completed by NK and verified by XJ for accuracy. Data was extracted into a predefined Microsoft Excel spreadsheet. A study ID was given to each included study. The extracted information included the study and participant characteristics (i.e., study design, country, sample size, gender), exposure details (SB and screen time description and measure), outcome details (MC description and measure) and key findings with respect to the association of sedentary behaviour or screen time with MC (including any co-variates included in the analysis). A p-value of <0.05 was used to denote statistically significant findings.

### Quality assessment for included studies

Quality of eligible studies was assessed independently by two reviewers (NK and JJR). Depending on the design of the study, the Joanna Briggs institute (JBI) critical appraisal tools were used on each study^11^. The JBI checklist was modified slightly-questions which referred to the standard criteria used for the measurement of the condition (disease), and whether participants were free from the outcome (disease) at the start of the study were not applicable to the review, hence, were not used in the assessment (see supplementary file 2). However, an additional question was included by authors to include assessment of the sample sizes used in the included studies, i.e., we assessed whether eligible studies considered sample size/ study power. This is because adequate sample size for a study provides an adequate power. Each category would receive a 1 if the criterion was met and a 0 of it was not. The sum of all categories was then calculated to create a total quality assessment score. The higher the score the higher the quality of the study. The adapted JBI critical appraisal checklist tool for the included studies is found in supplementary files 2 and 3.

### Synthesis of results

Due to the heterogeneity of the included studies, a meta-analysis could not be conducted. This was due to the differences in the measurement tools used in both the exposure and outcome measure. Instead, the vote counting method based on the direction of association was performed in line with the Cochrane Handbook for Systematic Reviews of Interventions^12^. Associations were classified as positive association where the exposure measure (screen time or habitual SB) were associated with higher or improved MC, while associations were classified as negative association if the exposure measure was associated with a lower MC (e.g., higher screen time usage associated with decreased MC). Results were summarised separately for the two exposure measures (screen time and habitual SB). The summary results were presented as the number of associations found divided by the total number of studies included. A binomial probability test was performed, the p-value of this test indicates the probability of observing the summary results if the exposure outcome associations were in the opposite direction^12^. This means the smaller the p-value, the higher the chances of the results being valid, independent of the p-values reported by the authors of the included studies.

## Results

### Description of studies

The database search yielded a total of 5276 studies of which 561 duplicates were removed. After screening of titles and abstracts 103 studies remained for full text screening. Following the full text screening, 16 studies were included for the final review (Figure 1). Excluded papers were studies that did not report on relevant associations (n = 56), included participants outside the age range (n = 22), were published outside the search time period (n = 4), were abstracts (n = 4) or a protocol paper (n = 1). Of the 16 included studies, 2 studies were longitudinal studies^13,22^, and 12 studies were cross-sectional^3,15,16,17,18,19,21,23,24,33,34,35^ with two studies reporting both cross-sectional and longitudinal results^14,20^. Six studies reported on time spent in habitual SB^,19,20,21,22,23,34^ as measured by accelerometer, seven studies reported on screen time^13,14,15,16,17,18,33^ as reported by parents using questionnaires and three studies reported on both SB and screen time ^3,24,35^. A summary of the selection and screening process is shown in the PRISMA flow diagram (Figure 1). Two of the included studies^17,22^ did not report a direction of association with the exposure and hence were not included in the vote counting results tables. Also, one study^3^ did not report a direction of association with habitual SB, balance, locomotor and object control subscales and hence was not included in the summary table.

**Fig 1.**
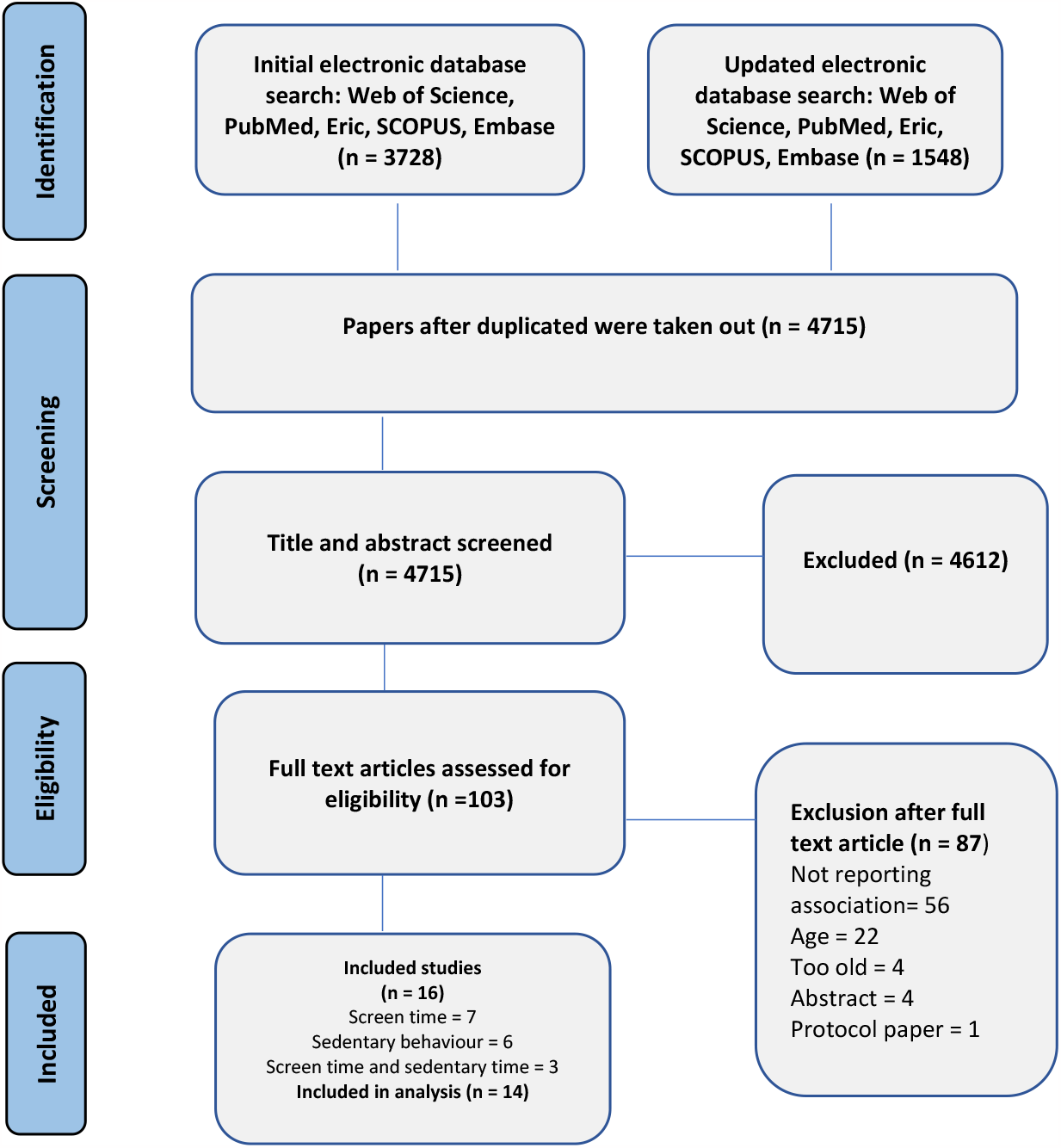
PRISMA Flow diagram of search and screening stages.

### Participants

The study samples for the 16 included studies included both boys and girls (total of 7,427 participants) from a range of different countries. (See Table 2).

**Table 2.**
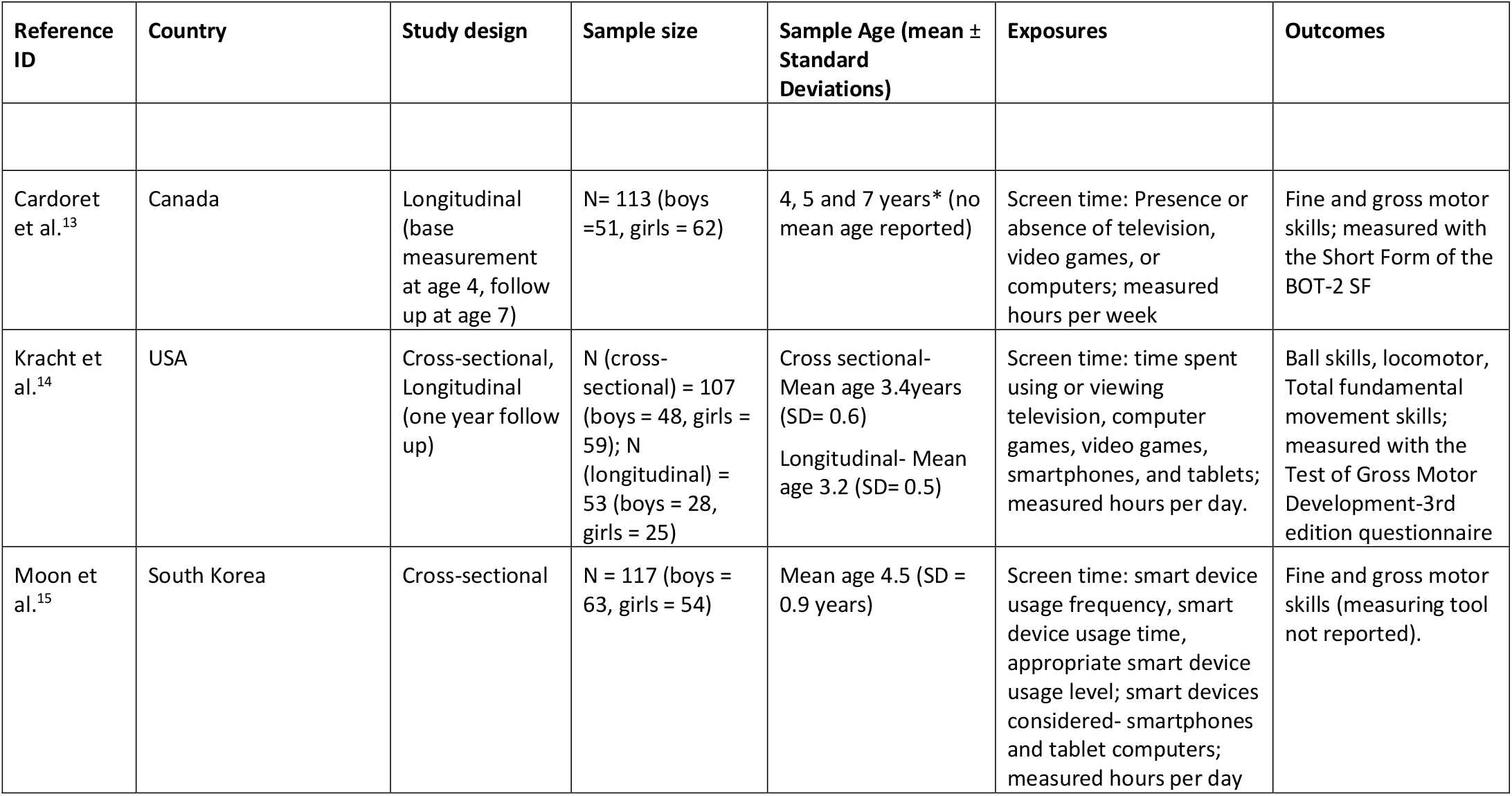
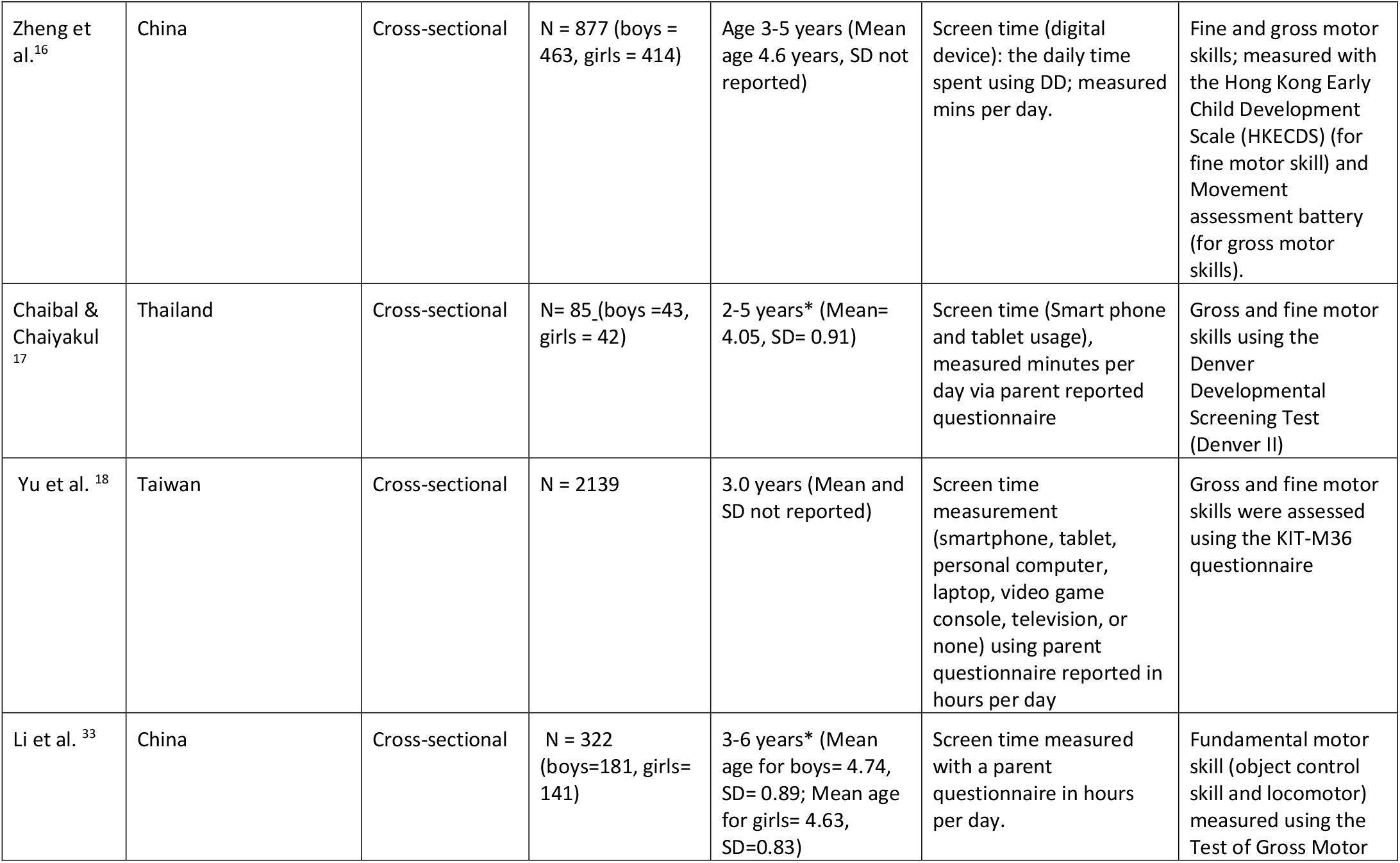
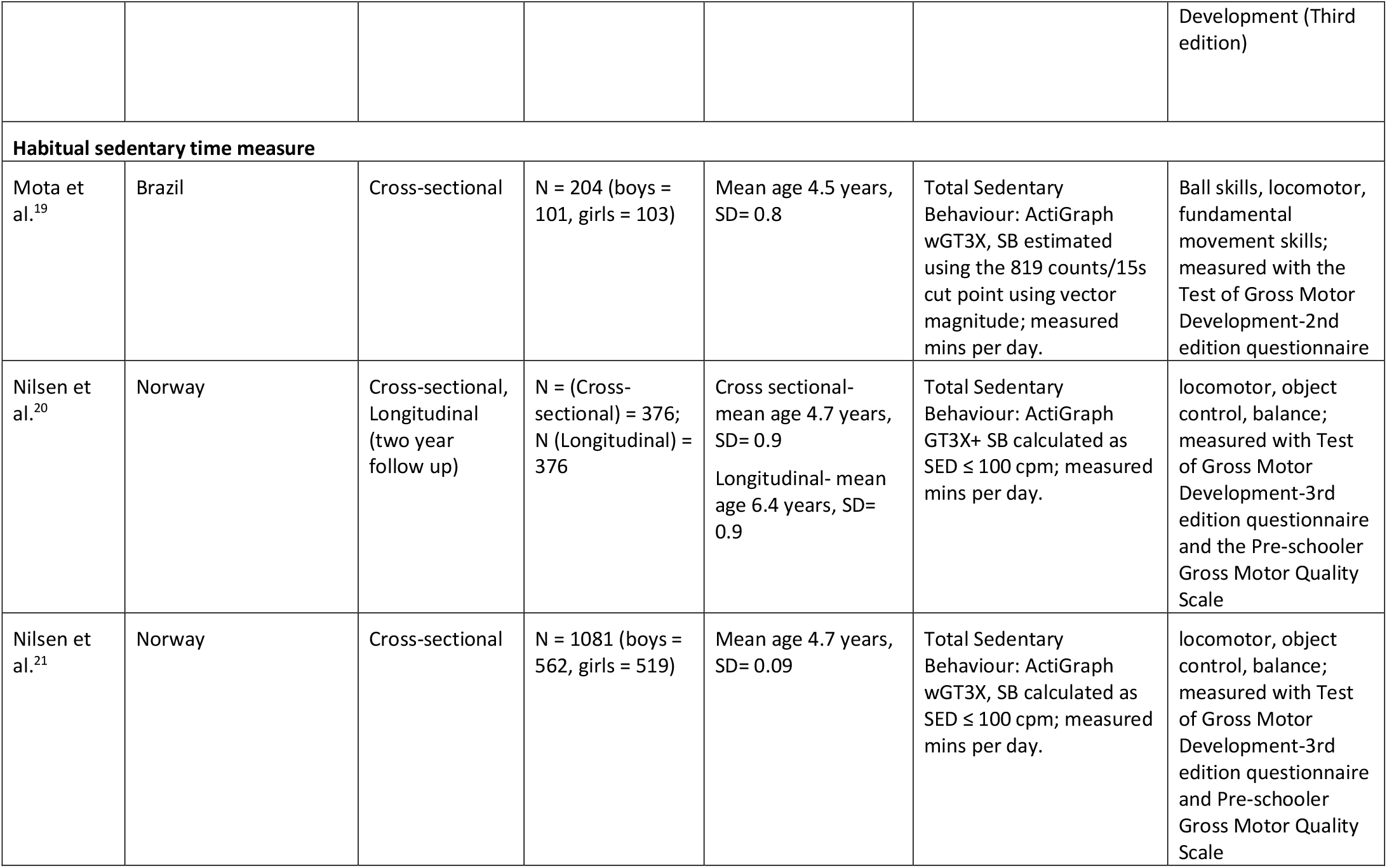
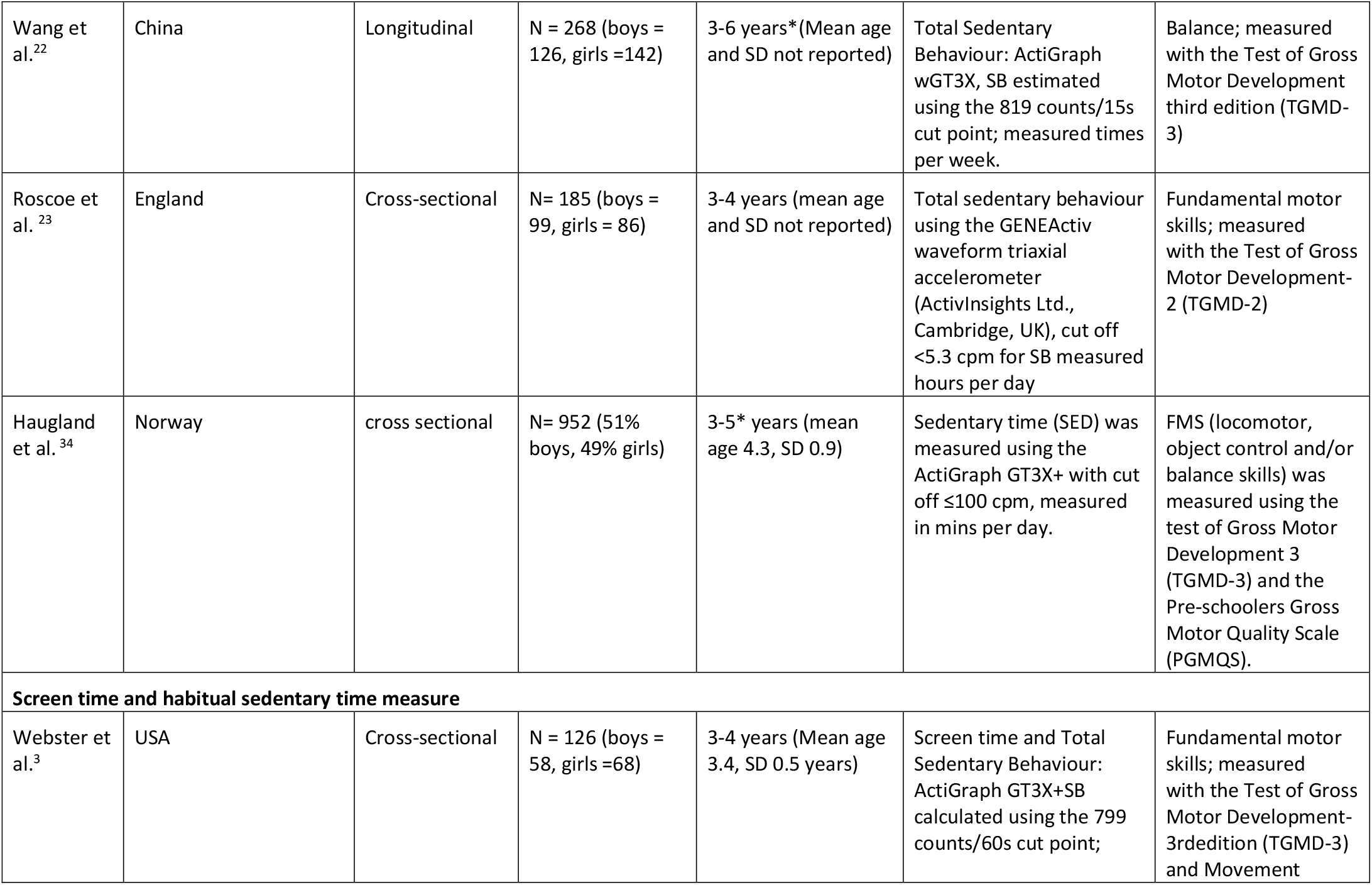
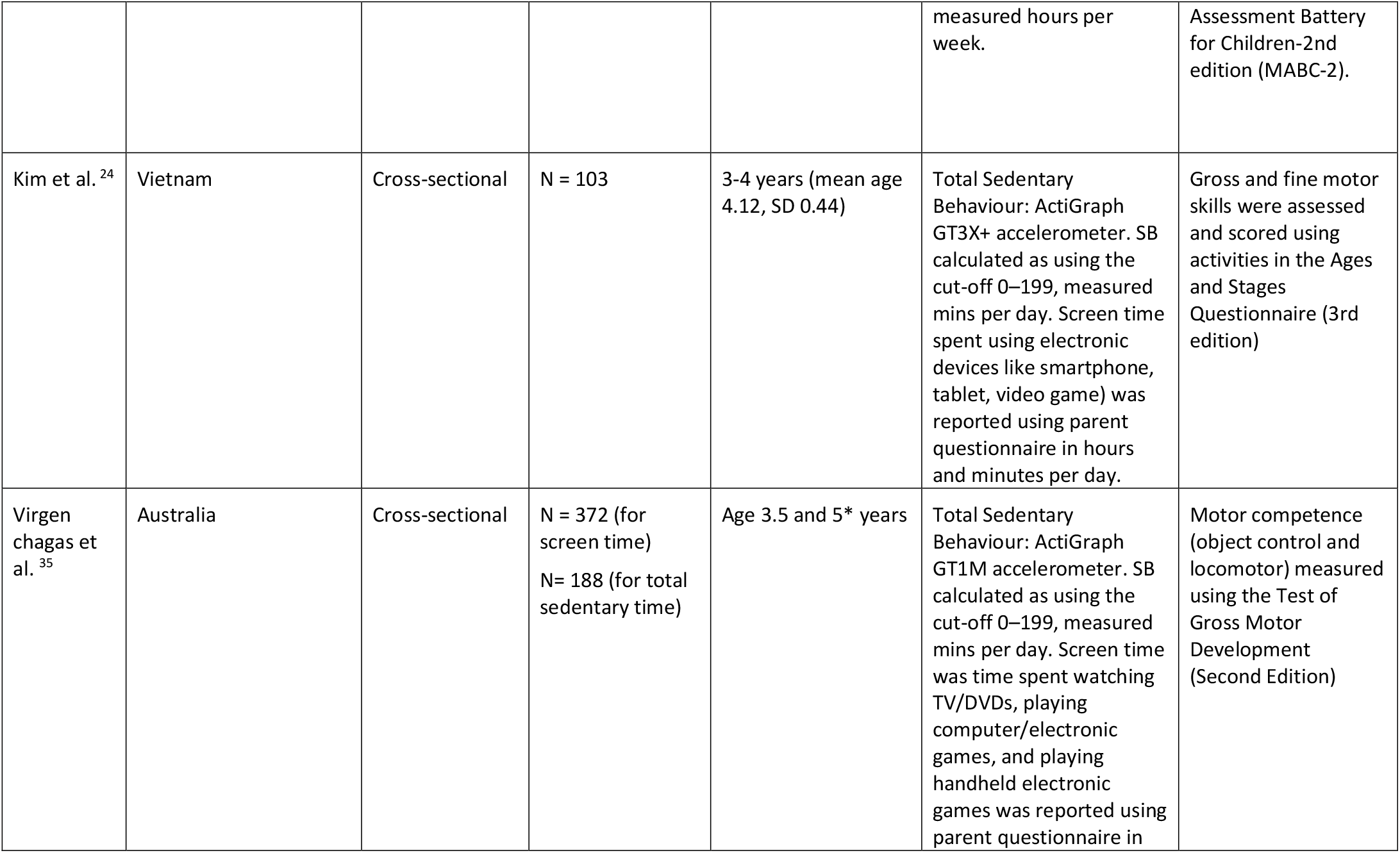
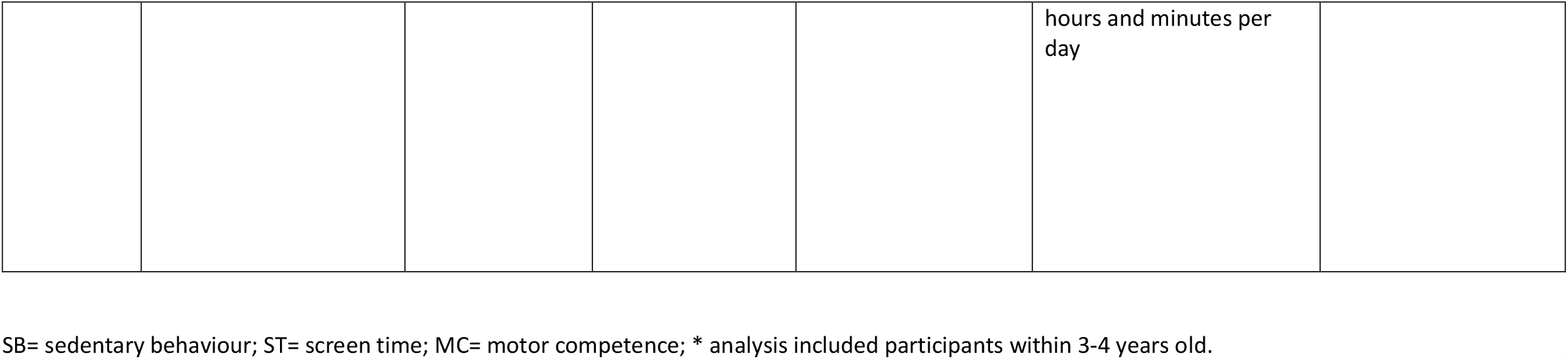
Descriptive characteristics of included studies.

| Reference ID | Country | Study design | Sample size | Sample Age (mean ± Standard Deviations) | Exposures | Outcomes |
| --- | --- | --- | --- | --- | --- | --- |
| Cardoret et al.<sup>13</sup> | Canada | Longitudinal (base measurement at age 4, follow up at age 7) | N= 113 (boys =51, girls = 62) | 4, 5 and 7 years* (no mean age reported) | Screen time: Presence or absence of television, video games, or computers; measured hours per week | Fine and gross motor skills; measured with the Short Form of the BOT-2 SF |
| Kracht et al.<sup>14</sup> | USA | Cross-sectional, Longitudinal (one year follow up) | N (cross-sectional) = 107 (boys = 48, girls = 59); N (longitudinal) = 53 (boys = 28, girls = 25) | Cross sectional- Mean age 3.4years (SD= 0.6)<br>Longitudinal- Mean age 3.2 (SD= 0.5) | Screen time: time spent using or viewing television, computer games, video games, smartphones, and tablets; measured hours per day. | Ball skills, locomotor, Total fundamental movement skills; measured with the Test of Gross Motor Development-3rd edition questionnaire |
| Moon et al.<sup>15</sup> | South Korea | Cross-sectional | N = 117 (boys = 63, girls = 54) | Mean age 4.5 (SD = 0.9 years) | Screen time: smart device usage frequency, smart device usage time, appropriate smart device usage level; smart devices considered- smartphones and tablet computers; measured hours per day | Fine and gross motor skills (measuring tool not reported). |
| Zheng et al.<sup>16</sup> | China | Cross-sectional | N = 877 (boys = 463, girls = 414) | Age 3-5 years (Mean age 4.6 years, SD not reported) | Screen time (digital device): the daily time spent using DD; measured mins per day. | Fine and gross motor skills; measured with the Hong Kong Early Child Development Scale (HKECDS) (for fine motor skill) and Movement assessment battery (for gross motor skills). |
| Chaibal & Chaiyakul<sup>17</sup> | Thailand | Cross-sectional | N= 85_(boys =43, girls = 42) | 2-5 years* (Mean= 4.05, SD= 0.91) | Screen time (Smart phone and tablet usage), measured minutes per day via parent reported questionnaire | Gross and fine motor skills using the Denver Developmental Screening Test (Denver II) |
| Yu et al.<sup>18</sup> | Taiwan | Cross-sectional | N = 2139 | 3.0 years (Mean and SD not reported) | Screen time measurement (smartphone, tablet, personal computer, laptop, video game console, television, or none) using parent questionnaire reported in hours per day | Gross and fine motor skills were assessed using the KIT-M36 questionnaire |
| Li et al.<sup>33</sup> | China | Cross-sectional | N = 322 (boys=181, girls= 141) | 3-6 years* (Mean age for boys= 4.74, SD= 0.89; Mean age for girls= 4.63, SD=0.83) | Screen time measured with a parent questionnaire in hours per day. | Fundamental motor skill (object control skill and locomotor) measured using the Test of Gross Motor |
|  |  |  |  |  |  | Development (Third edition) |
| **Habitual sedentary time measure** |  |  |  |  |  |  |
| Mota et al.<sup>19</sup> | Brazil | Cross-sectional | N = 204 (boys = 101, girls = 103) | Mean age 4.5 years, SD= 0.8 | Total Sedentary Behaviour: ActiGraph wGT3X, SB estimated using the 819 counts/15s cut point using vector magnitude; measured mins per day. | Ball skills, locomotor, fundamental movement skills; measured with the Test of Gross Motor Development-2nd edition questionnaire |
| Nilsen et al.<sup>20</sup> | Norway | Cross-sectional, Longitudinal (two year follow up) | N = (Cross-sectional) = 376; N (Longitudinal) = 376 | Cross sectional- mean age 4.7 years, SD= 0.9<br>Longitudinal- mean age 6.4 years, SD= 0.9 | Total Sedentary Behaviour: ActiGraph GT3X+ SB calculated as SED ≤ 100 cpm; measured mins per day. | locomotor, object control, balance; measured with Test of Gross Motor Development-3rd edition questionnaire and the Pre-schooler Gross Motor Quality Scale |
| Nilsen et al.<sup>21</sup> | Norway | Cross-sectional | N = 1081 (boys = 562, girls = 519) | Mean age 4.7 years, SD= 0.09 | Total Sedentary Behaviour: ActiGraph wGT3X, SB calculated as SED ≤ 100 cpm; measured mins per day. | locomotor, object control, balance; measured with Test of Gross Motor Development-3rd edition questionnaire and Pre-schooler Gross Motor Quality Scale |
| Wang et al.<sup>22</sup> | China | Longitudinal | N = 268 (boys = 126, girls =142) | 3-6 years*(Mean age and SD not reported) | Total Sedentary Behaviour: ActiGraph wGT3X, SB estimated using the 819 counts/15s cut point; measured times per week. | Balance; measured with the Test of Gross Motor Development third edition (TGMD-3) |
| Roscoe et al.<sup>23</sup> | England | Cross-sectional | N= 185 (boys = 99, girls = 86) | 3-4 years (mean age and SD not reported) | Total sedentary behaviour using the GENEActiv waveform triaxial accelerometer (ActivInsights Ltd., Cambridge, UK), cut off <5.3 cpm for SB measured hours per day | Fundamental motor skills; measured with the Test of Gross Motor Development-2 (TGMD-2) |
| Haugland et al.<sup>34</sup> | Norway | cross sectional | N= 952 (51% boys, 49% girls) | 3-5* years (mean age 4.3, SD 0.9) | Sedentary time (SED) was measured using the ActiGraph GT3X+ with cut off ≤100 cpm, measured in mins per day. | FMS (locomotor, object control and/or balance skills) was measured using the test of Gross Motor Development 3 (TGMD-3) and the Pre-schoolers Gross Motor Quality Scale (PGMQS). |
| **Screen time and habitual sedentary time measure** |  |  |  |  |  |  |
| Webster et al.<sup>3</sup> | USA | Cross-sectional | N = 126 (boys = 58, girls =68) | 3-4 years (Mean age 3.4, SD 0.5 years) | Screen time and Total Sedentary Behaviour: ActiGraph GT3X+SB calculated using the 799 counts/60s cut point; | Fundamental motor skills; measured with the Test of Gross Motor Development-3rd edition (TGMD-3) and Movement |
|  |  |  |  |  | measured hours per week. | Assessment Battery for Children-2nd edition (MABC-2). |
| Kim et al. <sup>24</sup> | Vietnam | Cross-sectional | N = 103 | 3-4 years (mean age 4.12, SD 0.44) | Total Sedentary Behaviour: ActiGraph GT3X+ accelerometer. SB calculated as using the cut-off 0–199, measured mins per day. Screen time spent using electronic devices like smartphone, tablet, video game) was reported using parent questionnaire in hours and minutes per day. | Gross and fine motor skills were assessed and scored using activities in the Ages and Stages Questionnaire (3rd edition) |
| Virgen chagas et al. <sup>35</sup> | Australia | Cross-sectional | N = 372 (for screen time)<br>N= 188 (for total sedentary time) | Age 3.5 and 5* years | Total Sedentary Behaviour: ActiGraph GT1M accelerometer. SB calculated as using the cut-off 0–199, measured mins per day. Screen time was time spent watching TV/DVDs, playing computer/electronic games, and playing handheld electronic games was reported using parent questionnaire in hours and minutes per day | Motor competence (object control and locomotor) measured using the Test of Gross Motor Development (Second Edition) |

SB= sedentary behaviour; ST= screen time; MC= motor competence; \* analysis included participants within 3-4 years old.

### Screen time

Associations between screen time and MC were examined in 10 of the included studies (Table 3). Of these 10, one was a longitudinal study, 8 were cross-sectional and 1 used both longitudinal and cross-sectional data. Screen time measures included parent-reported questionnaire to report time spent on tv, video games, computer games, smart phone/device usage, tablets and computers which were expressed in minutes per day, hours per day and hours per week. Outcome measures included fundamental motor skills, gross and fine motor skill, locomotor skill, object control skills, measured with the Test of Gross Motor Development questionnaire 2nd and 3rd edition, the Short Form of the BOT-2 SF, the Hong Kong Early Child Development Scale (HKECDS) (for fine motor skill), the Movement assessment battery and the Denver Developmental Screening Test (Denver II).

**Table 3.** Summary table of exposure (screen time) and outcome associations.

| Exposure: Screen time |  |  |  |  |  |
| --- | --- | --- | --- | --- | --- |
| Outcome | Negatively associated to exposure <sup>a</sup> | Positively associated to exposure <sup>a</sup> | Summary |  | N participants (Total or range) |
|  |  |  | n/N (%) | P value |  |
| Fundamental motor skills | 3, **13, 14, 18** | 14 | 4/5 (80) | 0.19 | cross sectional (107-126); longitudinal (166) |
| Gross motor skill | 15*, **16**, 18, 24 | 24, 15** | 4/6 (67) | 0.34 | cross sectional (877- 926) |
| Fine motor skills | **3**, 15*, 15** 16, **18**, 24 | 24 | 6/7 (86) | 0.06 | cross sectional (877) |
| Locomotor skill | 14, 35 | 14, 33 | 2/4 (50) | 0.69 | Cross sectional (107-372)<br>Longitudinal (53) |
| Object control | 14, 14, 33 | 35 | 3/4 (75) | 0.31 | cross sectional (107-372)<br>Longitudinal (53) |

n = number of negative associations; N = total number of associations for the exposure outcome relation. <sup>a</sup> = numbers refer to reference numbers. Studies that found significant associations in bold. *Longitudinal studies in italics*. \*Negative associations in 3yr olds with exposure, \*\* positive association in 4yr olds with exposure.

Mixed results were reported for fundamental motor skills (4 out of 5 negative associations), gross motor skills (4 out of 6 negative associations), fine motor skills (6 out of 7 negative associations), locomotor skill (2 out of 4 negative associations) and object control (3 out of 4 negative associations).

Significant associations were only found in those studies reporting negative associations between screen time and MC ^3,13,14,16,18^.

Two longitudinal studies found a negative association with FMS^13,14^ and one with locomotor skill^14^ and object control^14^. For cross sectional studies, there were negative associations with FMS^3,18^, gross motor skill^15,16,18,24^, fine motor skills^,3,15,16,18,24^, locomotor^35^ and object control^14,33^.

### Habitual sedentary behaviour

Associations between time spent in SB and MC were examined in 9 of the included studies (Table 4). Of these 9 studies, 1 was longitudinal, 7 were cross-sectional and 1 used both longitudinal and cross-sectional data. Habitual SB was measured using the ActiGraph GT3X+ and the GENEActiv waveform triaxial accelerometer. SB was reported as minutes per day, hours per day and times per week. Outcome measure included fundamental motor skills, gross and fine motor skill, locomotor skill, object control skill and balance, measured with the Test of Gross Motor Development questionnaire 2nd and 3rd edition), the Movement Assessment Battery for Children-2nd edition (MABC-2) and Pre-schooler Gross Motor Quality Scale.

**Table 4.** Summary table of exposure (sedentary behaviour) and outcome associations.

| Exposure: Habitual sedentary behaviour |  |  |  |  |  |
| --- | --- | --- | --- | --- | --- |
| Outcome | Negatively associated to exposure <sup>a</sup> | Positively associated to exposure <sup>a</sup> | Summary |  | N participants (Total or range) |
|  |  |  | n/N (%) | P value |  |
| Fundamental motor skills | 19<sup>*</sup>, 23<sup>*</sup> weekend, **23<sup>\*\*</sup>** weekdays, **23<sup>\*\*</sup>** 4days | **19<sup>\*\*</sup>**, **23<sup>\*</sup>** week, 23<sup>*</sup> 4days, **23<sup>\*\*</sup>** weekend | 4/8 (50) | 0.66 | cross sectional (185-204) |
| Locomotor skill | 19<sup>*</sup>, 23<sup>*</sup> weekend, 23<sup>*</sup> 4days, **23<sup>\*\*</sup>** weekdays, **23<sup>\*\*</sup>** 4days **20, 20, 21, 34, 35** | 19<sup>**</sup>, 23<sup>*</sup> week, **23<sup>\*\*</sup>** weekend | 10/13 (73) | 0.05 | cross sectional (185-1081), longitudinal (224) |
| Object control | 19<sup>*</sup>, 23<sup>*</sup> weekend, **23<sup>\*\*</sup>** weekdays, **23<sup>\*\*</sup>** 4days **20, 20, 21, \* 34, 35** | **19<sup>\*\*</sup>**, 23<sup>*</sup> 4days, **23<sup>\*</sup>** week, **23<sup>\*\*</sup>** weekend | 9/13 (64) | 0.13 | Cross sectional (185-1081) longitudinal (224) |
| Balance | 20, 20, 21, 34 |  | 4/4 (100) | 0.06 | Cross sectional (217-1081) longitudinal (224) |

n = number of negative associations; N = total number of associations for the exposure outcome relation. <sup>a</sup> = numbers refer to reference numbers. Studies that found significant associations in bold. *Longitudinal studies in italics*. \*Sedentary behaviour is replaced with moderate-to-vigorous physical activity; \*\*sedentary behaviour is replaced by sleep or light physical activity.

Negative associations were reported for balance (4/4). Mixed associations were reported for fundamental motor skills (4 out of 8 negative associations), locomotor skill (10 out of 13 negative associations), and object control (9 out of 13 negative associations). Significant associations were found in those studies reporting both negative associations between SB and MC^20,21,23^ and positive association between SB and MC^19,23^. For habitual SB time, longitudinal studies found a negative association with locomotor skill^20^, object control^20^ and balance^20^. For cross sectional studies, there were negative associations with FMS^19,23^ locomotor skill^19,20,21,23,34,35^, object control^19,20,21,23,34,35^ and balance^20,21,34^.

### Quality assessment

The overall quality of evidence for the study scores ranged from 3 to 7 (out of a potential 8) for the cross-sectional studies and 6 to 9 (out of potential 11) for the longitudinal studies (Tables 4 and 5). The main source of potential bias was sample size. No studies provided a justification of the sample size used for the research. Another source of bias included missing information on the validity and reliability of some of the measurement tool used for the exposure variables (sedentary behaviour including screen time).

**Table 5.** Quality assessment of included cross-sectional studies ^11^.

| Study | Inclusion criteria | Subjects and setting detailed | Valid/reliable exposure measure | measure of condition | Confounding factors identified | Strategy for confounding factors stated | Valid/reliable outcome measure | Appropriate statistical analysis | Adequate sample size | Total Score |
| --- | --- | --- | --- | --- | --- | --- | --- | --- | --- | --- |
| Mota et al. <sup>19</sup> | Y | Y | Y | N/A | U | U | Y | Y | N | 5 |
| Moon et al. <sup>15</sup> | Y | Y | U | N/A | N | N | U | Y | N | 3 |
| Zheng et al. <sup>16</sup> | Y | Y | U | N/A | Y | Y | Y | Y | N | 6 |
| Wang et al. <sup>22</sup> | Y | Y | Y | N/A | U | U | Y | Y | N | 5 |
| Webster et al. <sup>3</sup> | Y | Y | U | N/A | Y | Y | Y | Y | N | 6 |
| Chaibal & Chaiyakul. <sup>17</sup> | Y | Y | N | N/A | Y | Y | Y | Y | N | 6 |
| Yu et al. <sup>18</sup> | Y | Y | N | N/A | U | U | N | U | Y | 3 |
| Roscoe et al. <sup>23</sup> | Y | Y | U | N/A | U | U | Y | Y | N | 4 |
| Kim et al. <sup>24</sup> | Y | Y | U | N/A | Y | Y | Y | Y | N | 6 |
| Haugland et al. <sup>34</sup> | Y | Y | Y | N/A | Y | Y | Y | Y | N | 7 |
| Li et al. <sup>33</sup> | Y | Y | U* | N/A | Y | Y | Y | Y | N | 6 |
| Virgen Chagas et al. <sup>35</sup> | Y | Y | U | N/A | U | U | Y | Y | N | 4 |

Y = Yes, U = unclear, N = No, N/A = Not applicable, U\* = accelerometry measure valid and reliable, parent questionnaire unclear of validity/reliability.

**Table 6.** Quality assessment of included longitudinal studies^11^.

| **Study** | **Inclusion criteria** | **Measure** | **validity and reliability of exposure** | **Confounding factors identified** | **Strategy for confounding factors stated** | **participants free from outcome** | **validity and reliability of outcome measure** | **Reported follow up** | **follow up described and explored** | **strategies for incomplete follow ups** | **appropriate statistical analysis** | **sample size** | **Total score** |
| --- | --- | --- | --- | --- | --- | --- | --- | --- | --- | --- | --- | --- | --- |
| Cardoret et al.<sup>13</sup> | Y | Y | Y | N | N | N/A | Y | Y | U | U | Y | N | **6** |
| Kracht et al.<sup>14</sup> | Y | Y | U | Y | Y | N/A | Y | Y | N | N | Y | N | **7** |
| Nilsen et al.<sup>20</sup> | Y | Y | Y | Y | Y | N/A | Y | Y | Y | N | Y | N | **9** |
| Nilsen et al.<sup>21</sup> | Y | Y | Y | Y | Y | N/A | Y | Y | N | N | Y | N | **8** |

Y = Yes, U = unclear, N = No, N/A = Not applicable

## Discussion

The purpose of this study was to conduct a systematic review of recent studies to examine the associations between time spent in different components of both SB (screen time and habitual SB) and MC (fundamental motor skills, fine and gross motor skills, locomotor skills, ball skills/object control and balance) among 3-4 -year-olds. For this review, screen time was time spent on electronic devices (watching tv, video games, time spent on tablets/phones) which was generally reported via parent questionnaire, while habitual sedentary behaviour was any posture with an energy expenditure ≤1.5 metabolic equivalents of task (MET) whiles awake (e.g., sitting, lying, time spent in a buggy) and was generally reported with the use of an ActiGraph accelerometer. Findings from this review shows predominantly negative associations between both screen time and time spent in habitual SB with different components of MC (FMS, fine and gross motor skills, locomotor skills, object control and balance). For instance, for screen time measurement there was a predominantly negative association with fundamental motor skill (4 out of 5 studies), fine motor skill (6 out of 7 studies), gross motor skill (4 out of 6 studies) and object control (3 out of 4 studies). For habitual sedentary behaviour time, there was also a predominantly negative association with locomotor skill (10 out of 13 studies) and object control (9 out of 13 studies), with four studies all reporting negative association with balance (4 out of 4 studies). These associations however did not show a statistical significance due to fewer studies used for the analysis which highlights a limitation with the analytical method used. With the method of vote counting, a binomial probability test was conducted and p-values from this test indicated the probability of observing the summary results if the exposure and outcome associations were in the opposite direction. This method usually focuses on the direction of association as the p-values from this test might be affected by the number of studies used for the analyses. Time spent on screen time and habitual SB may therefore unfavourably affect the development of motor competence, which is crucial for physical activities later in life.

Findings from this review expand on the previous review by Poitras et al.^8^ which concluded that the associations between screen time and MC from studies up to April 2016 were largely unfavourable/null. Poitras et al. ^8^ found that for total sedentary behaviour, one study showed no association with motor skills at age 3-4 years and another also showing a % sedentary time being negatively associated with locomotor skill at age 3-4 years. For screen-based SB, Poitras et al. ^8^ also found TV time to be negatively associated with motor skill development.

The review by Dos Santos et al. ^9^ also found generally negative associations between time spent in SB and MC among children and adolescents. Although findings from this review is similar to the review by Dos Santos et al. considered 3-6-year-olds together, while this current review included children between 3 and 4 years in line with the WHO age group for early years. Also, Dos Santos et al. did not differentiate between screen time and habitual SB or the different components of MC (FMS, fine and gross motor skills, locomotor skills, ball skills/object control and balance)-this current review did.

Recent research has reported that only a small proportion of pre-schoolers between the ages of 3 and 4 years adhere to the sedentary behaviour guidelines by the WHO, implying that excess screen time reduces time spent on physical activity and/or sleep in the majority of children since in a fixed 24 hour period higher screen time must result in lower time spent in physical activity and/or sleep^25,26,27^. The level of MC in the early years not only influences physical activity, but also other health indicators like cardiorespiratory fitness and adiposity^27,28^. Children who spend more time in SB may have fewer opportunities to develop their MC^9,29^. Therefore, reducing the time a child spends sedentary may assist the acquisition of adequate MC which can in its turn influence physical activity and health later in life^6,27^.

Some strengths of the current review include the application of rigorous methodological standards established for conducting a systematic review (PRISMA^30^). Authors used a comprehensive search strategy developed with the help of a librarian with an expertise in systematic review and searched a comprehensive list of the most relevant databases. The quality of the included studies was assessed using the Joanna Briggs Institute assessment tools which are both valid and reliable^11^. The review assessed the individual components of both times spent in habitual SB and motor competence to provide a greater understanding on the impact different SB might have on different aspects of MC. The review does not come without limitations. Studies were limited to those published in English language only. Also due to the heterogeneous nature in the exposure and outcome measure, meta-analysis was not possible. Finally, the majority of studies included small sample sizes (10 out of 13 eligible studies had fewer than 500 participants), this may have limited their statistical power. Hence, using the method of vote counting, which is based on the direction of association, may have limited the impact of the underpowered studies in the summarized results^13^.

This review has also highlighted several areas for future research. Most of the included studies used parent questionnaires for measurement of screen time-validity and reliability of these has not been ascertained within 3-4-year-olds^31^. There is a need for the development of proxy reported or other instruments that can accurately capture screen time. Establishing a high-quality standard measure for time spent in habitual SB remains a challenge, and a uniform approach to data collection would help minimize some of the challenges associated with quantifying SB in young children as well as providing a more comparable evidence base. In future studies, measuring tools such as inclinometers which capture postures more accurately than the accelerometer^32^, or limb worn devices can help with some of the challenges associated with quantifying habitual SB in early years. Further studies with a stronger study design (such as prospective studies and intervention studies) will be required to confirm the associations between screen time and/or SB and MC in 3–4-year-olds.

## Conclusion

This systematic review synthesized findings from 16 studies with 7,424 participants from different parts of the world. Quality of evidence was moderate. In summary this review showed a predominantly negative association of screen time and habitual SB with motor competence in 3- and 4-year-olds. Findings from this review extend and support the findings of previous reviews and highlight the importance of limiting both screen time and habitual SB in early years for prevention of disease and optimal promotion of health. Future research that uses stronger study designs (e.g., longitudinal or retrospective studies) as well as valid and reliable measurement tool is required to validate findings from this review.

## Data Availability

All data produced in the present work are contained in the manuscript

## Key messages

- Targeting high levels of screen time and sedentary behaviour may be used as a strategy to increase motor competence in young children.
- Studies examining the different types of sedentary behaviour on different components of motor competence are needed.
- High-quality studies with strong designs are needed to strengthen the evidence base.
- There is need for the development of self-reported or other instruments that can accurately capture screen time measurement and can do so across different screen-based devices.

## Search strategy

An electronic literature search was conducted on the following databases on the 27^th^ of July 2021 (updated on 30^th^ September 2023): Web of Science core collection, PUBMED Central, EMBASE (Ovid), and SPORT Discus and ERIC. For each database search, keywords and synonyms were developed using the PICO (Populations, Intervention/exposure, Comparator and Outcome) framework. Keywords were identified and used in each database with specific database functions and tools utilised.

Summary of Keywords and combination used for the database search include

1. “Early childhood” or “Preschool children” or “Pre-school children” or “early years” or “children” (population)
2. “sedentary” or “Sedentary behaviour” or “sedentary lifestyle” or “sedentary activities” or “physical inactivity” or “inactivity” or “screen time” or “screen based” or “sitting time” or “television time” or “screen-based entertainment” or “computer” or “video games” or “smartphone” (intervention/exposure)
3. “Motor development” or “Motor Skill” or “motor competence” or “fundamental motor skill” or “motor proficiency” or “psychomotor performance” or “fine motor skill” or “gross motor skill” or “locomotor control” or “object control” or “child development” or “growth and development” or “ball skills” or “stability” or “balance” (primary outcomes)
4. 1 and 2 and 3

## PubMed Central

Notes:

“Early childhood” or “Preschool children” or “Pre-school children” or “early years” or “children” (population)

2. “sedentary” or “Sedentary behaviour” or “sedentary lifestyle” or “sedentary activities” or “physical inactivity” or “inactivity” or “screen time” or “screen based” or “sitting time” or “television time” or “screen-based entertainment” or “computer” or “video games” or “smartphone” (intervention/exposure)
3. “Motor development” or “Motor Skill” or “motor competence” or “fundamental motor skill” or “motor proficiency” or “psychomotor performance” or “fine motor skill” or “gross motor skill” or “locomotor control” or “object control” or “child development” or “growth and development” or “ball skills” or “stability” or “balance” (primary outcomes)
4. 1 and 2 and 3

## Sport Discus

Notes:

- Abstract search field selected
  1. “Early childhood” or “Preschool children” or “Pre-school children” or “early years” or “children” (population)
  2. “sedentary” or “Sedentary behaviour” or “sedentary lifestyle” or “sedentary activities” or “physical inactivity” or “inactivity” or “screen time” or “screen based” or “sitting time” or “television time” or “screen-based entertainment” or “computer” or “video games” or “smartphone” (intervention/exposure)
  3. “Motor development” or “Motor Skill” or “motor competence” or “fundamental motor skill” or “motor proficiency” or “psychomotor performance” or “fine motor skill” or “gross motor skill” or “locomotor control” or “object control” or “child development” or “growth and development” or “ball skills” or “stability” or “balance” (primary outcomes)
  4. 1 and 2 and 3

## ERIC

Notes:

- Abstract search field selected

## Web of Science: Core collection

Notes

- Topic search field was selected.
  1. “Early childhood” or “Preschool children” or “Pre-school children” or “early years” or “children” (population)
  2. “sedentary” or “Sedentary behaviour” or “sedentary lifestyle” or “sedentary activities” or “physical inactivity” or “inactivity” or “screen time” or “screen based” or “sitting time” or “television time” or “screen-based entertainment” or “computer” or “video games” or “smartphone” (intervention/exposure)
  3. “Motor development” or “Motor Skill” or “motor competence” or “fundamental motor skill” or “motor proficiency” or “psychomotor performance” or “fine motor skill” or “gross motor skill” or “locomotor control” or “object control” or “child development” or “growth and development” or “ball skills” or “stability” or “balance” (primary outcomes)
  4. 1 and 2 and 3

## EMBASE: Excerpta Medica (Ovid)

Notes:

